# Rapid implementation of mobile technology for real-time epidemiology of COVID-19

**DOI:** 10.1101/2020.04.02.20051334

**Authors:** David A. Drew, Long H. Nguyen, Claire J. Steves, Jonathan Wolf, Tim D. Spector, Andrew T. Chan, on behalf of the COPE Consortium

## Abstract

The rapid pace of the severe acute respiratory syndrome coronavirus 2 (SARS-CoV-2) pandemic (COVID-19) presents challenges to the robust collection of population-scale data to address this global health crisis. We established the COronavirus Pandemic Epidemiology (COPE) consortium to bring together scientists with expertise in big data research and epidemiology to develop a COVID-19 Symptom Tracker mobile application that we launched in the UK on March 24, 2020 and the US on March 29, 2020 garnering more than 2.25 million users to date. This mobile application offers data on risk factors, herald symptoms, clinical outcomes, and geographical hot spots. This initiative offers critical proof-of-concept for the repurposing of existing approaches to enable rapidly scalable epidemiologic data collection and analysis which is critical for a data-driven response to this public health challenge.

**One Sentence Summary:** COVID-19 symptom tracker for smartphones

## Main Text

The exponentially increasing number of severe acute respiratory syndrome coronavirus 2 (SARS-CoV-2) infections has led to “an urgent need to expand public health activities to elucidate the epidemiology of the novel virus and characterize it’s potential impact.”(*1*) Understanding risk factors for infection and predictors of subsequent outcomes is critical to gain control of the coronavirus disease 2019 (COVID-19) pandemic (*2*). However, the speed at which the pandemic is unfolding poses an unprecedented challenge to collecting exposure data characterizing the full breadth of disease severity, hampering efforts to disseminate accurate information in a timely manner to impact public health planning and clinical management. Thus, there is an urgent need for an adaptable real-time data-capture platform to rapidly and prospectively collect actionable high-quality data that encompasses the spectrum of subclinical and acute presentations while identifying disparities in diagnosis, treatment, and clinical outcomes. Addressing this priority will allow for more accurate estimates of disease incidence, inform risk mitigation strategies, more effectively allocate still-scarce testing resources, and allow for appropriate quarantine and treatment of those afflicted.

An evolving body of literature suggests COVID-19 incidence and outcomes vary according to age, sex, race/ethnicity, and underlying health status, with inconsistent evidence suggesting that commonly used medications such as angiotensin-converting enzyme (ACE) inhibitors, thiazolidinediones (TZD), and ibuprofen may alter the natural disease course(*3-9*). Further, symptoms of COVID-19 vary widely, with fever and dry cough reportedly the most prevalent, though numerous investigations have demonstrated that asymptomatic carriage is a significant determinant of community spread(*3, 4, 6, 10-14*). In addition, the full spectrum of clinical presentation is still being characterized, which may be significantly different in different patient groups, as evidenced by recent advisories by the American Gastroenterological Association (AGA) and the American Academy of Otolaryngology - Head and Neck Surgery (AAO-HNS), and British Geriatric Society (BGS) on the potential importance of previously underappreciated gastrointestinal symptoms (e.g. nausea, anorexia, and diarrhea) or loss of taste and/or smell associated with COVID-19 infection, as well as common geriatric syndromes e.g. falls and delirium. The pandemic has dramatically outpaced our collective efforts to fully characterize who is most at-risk and who may suffer the most serious sequelae of infection.

Mobile phone applications or web-based tools facilitate self-guided collection of population-level data at scale(*15*), the results of which can then be rapidly redeployed to inform participants of urgent health information (*15, 16*). Both are particularly advantageous when more than three-quarters of Americans are advised to physically distance(*13*). Such digital tools have already been used in more controlled research settings which benefit from greater lead time for field testing, question curation, and recruitment. Although several digital collection tools for COVID-19 symptoms have been developed and launched in the U.S., including some in partnership with government health agencies such as the CDC, these applications have largely been configured to offer a single assessment of symptoms to tailor recommendations for further evaluation. Alternatively, others have been developed for researchers to report patient-level information on behalf of participants already enrolled in clinical registries. While these approaches offer critical public health insights, they are not tailored for the type of scalable longitudinal data capture that epidemiologists need to perform comprehensive, well-powered investigations to address this public health crisis.

To meet this challenge, we established an international collaboration, the COronavirus Pandemic Epidemiology (COPE) consortium, comprised of leading investigators from several large clinical and epidemiological cohort studies. COPE brings together a multidisciplinary team of scientists with expertise in big data research and translational epidemiology to interrogate the COVID-19 pandemic in the largest and most diverse patient population assembled to-date. Several large inception cohorts have already agreed to join these efforts, including the Nurses’ Health Study II and 3, the Growing Up Today Study, the Health Professionals Follow-Up Study, TwinsUK, Cancer Prevention Study 3, and the Multiethnic Cohort. To aid in our data harmonization efforts, we co-developed a COVID-19 Symptom Tracker mobile app in partnership with in-kind contributions from Zoe Global Ltd, a digital healthcare company and academic scientists from King’s College London. By leveraging the established digital backbone of an application used for personal nutrition studies, the COVID Symptom Tracker was launched in the UK on March 24, 2020, garnering 1,9,000 users over 8 days since launch. We launched the app in the U.S. on the evening of March 29th, 2020.

The COVID Symptom Tracker enables self-report of data related to COVID-19 exposure and infections (Fig. 1). On first use, the app queries location, age, and core health risk factors. Daily prompts query for updates on interim symptoms, health care visits, and COVID-19 testing results. In those self-quarantining or seeking health care, the level of intervention and related outcomes are collected. Individuals without obvious symptoms are also encouraged to use the app. Through pushed software updates, we can add or modify questions in real-time to test emerging hypotheses about COVID-19 symptoms and treatments. Importantly, participants enrolled in ongoing epidemiologic studies, clinical cohorts, or clinical trials can provide informed consent to link survey data collected through the app in a Health Insurance Portability and Accountability Act (HIPPA)- and General Data Protection Regulation (GDPR)-compliant manner to their pre-existing study cohort data and any relevant biospecimens. A specific module is also provided for participants who identify as healthcare workers to determine the intensity and type of their direct patient care experiences, the availability and use of personal protective equipment (PPE), and work-related stress and anxiety.

**Fig. 1.**
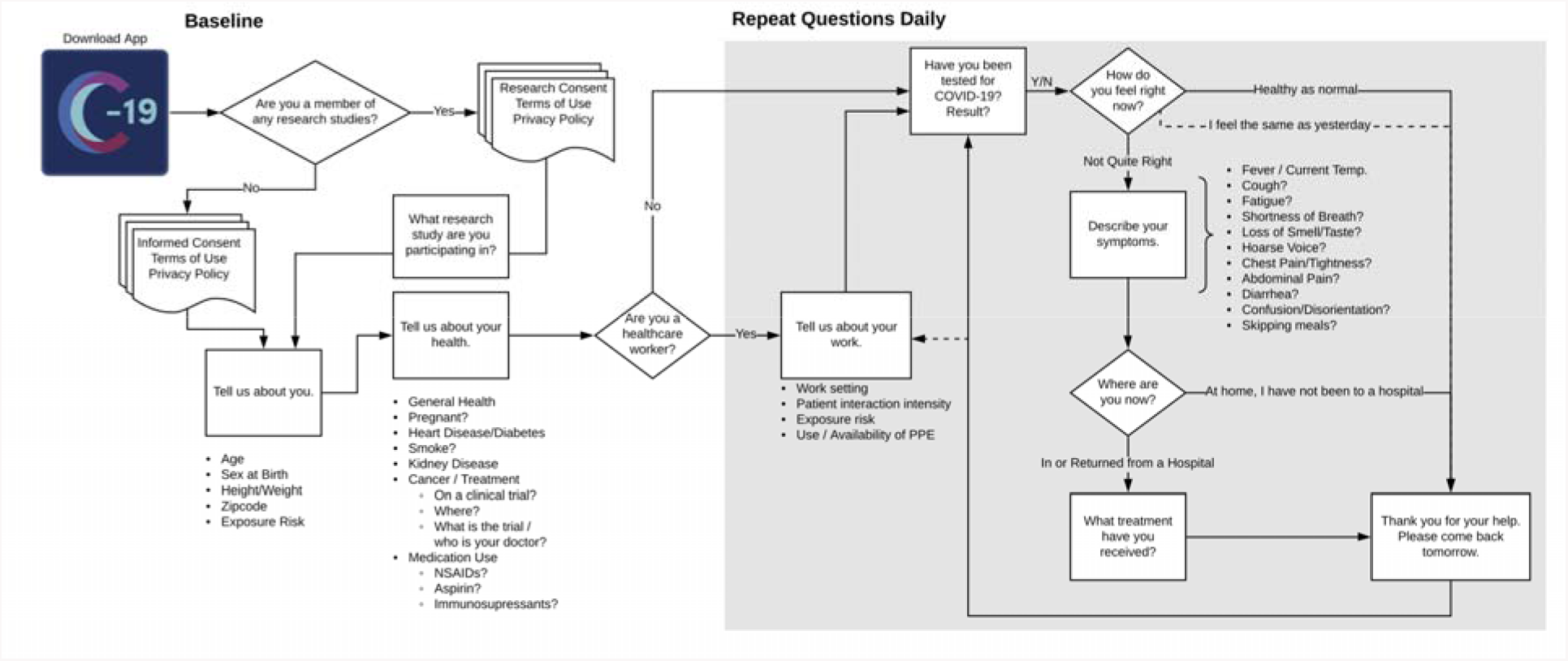
A schematic of participant workflow. After downloading the COVID Symptom Tracker and providing consent, users are prompted to provide baseline demographic and clinical information and are serially queried about new or ongoing symptoms, Health care workers offer additional information about the intensity of their patient interactions, potential exposure to infected patients, and usage of personal protective equipment. With informed consent, users already engaging in active research as participants in a variety of ongoing cohorts or clinical trials (i.e. Nurses’ Health Study, TwinsUK, etc.) have the option of linking COVID Symptom Tracker information to their extant research data.

Through rapid deployment of this tool, we can gain critical insights into population dynamics of the disease (Fig. 2). By collecting participant-reported geospatial data, highlighted as a critical need for pandemic epidemiologic research (*16*), we can rapidly identify populations with highly prevalent symptoms that may emerge as hot spots for outbreaks. A preliminary snapshot of the first 1.6 million users in the UK over the first five days of use confirms the variability in symptoms reported across suspected COVID-19 cases and is useful for generating and testing broader hypotheses. Users are a mean age of 41 with a range from 18 to 90 years, with 75% female users. Simple visualization of initial results (Fig. 3) demonstrates that among those reporting symptoms by March 27, 2020 (n=265,851) the most common symptoms were fatigue and cough, followed by diarrhea, fever, and anosmia. Shortness of breath was relatively rarely reported. Only 0.2% (n=744) of individuals reporting possible COVID-19 symptoms reported receiving a qPCR test for COVID-19. Among individuals who did undergo a test, cough and fatigue alone or in combination commonly led to testing, but was not highly predictive of a positive test. Similarly, no individuals reporting diarrhea in the absence of other symptoms tested positive. Interestingly, more complex presentations with cough and/or fatigue and at least one additional symptom, including less commonly appreciated symptoms such as diarrhea and anosmia, appeared to be better predictors. In particular, anosmia may be a more sensitive symptom as it was more common than fever in individuals who tested positive. In contrast, fever alone was not particularly discriminatory. However, in combination with lesser appreciated symptoms, a greater frequency of positive tests was observed. These findings suggest that individuals with complex symptomatic presentation perhaps should be prioritized for testing. Concerningly, 20% of individuals report complex symptoms (cough and/or fatigue plus at least one of anosmia, diarrhea, or fever) but have not yet received testing, representing a substantial population who appear to be at greater risk for the disease.

**Fig. 2.**
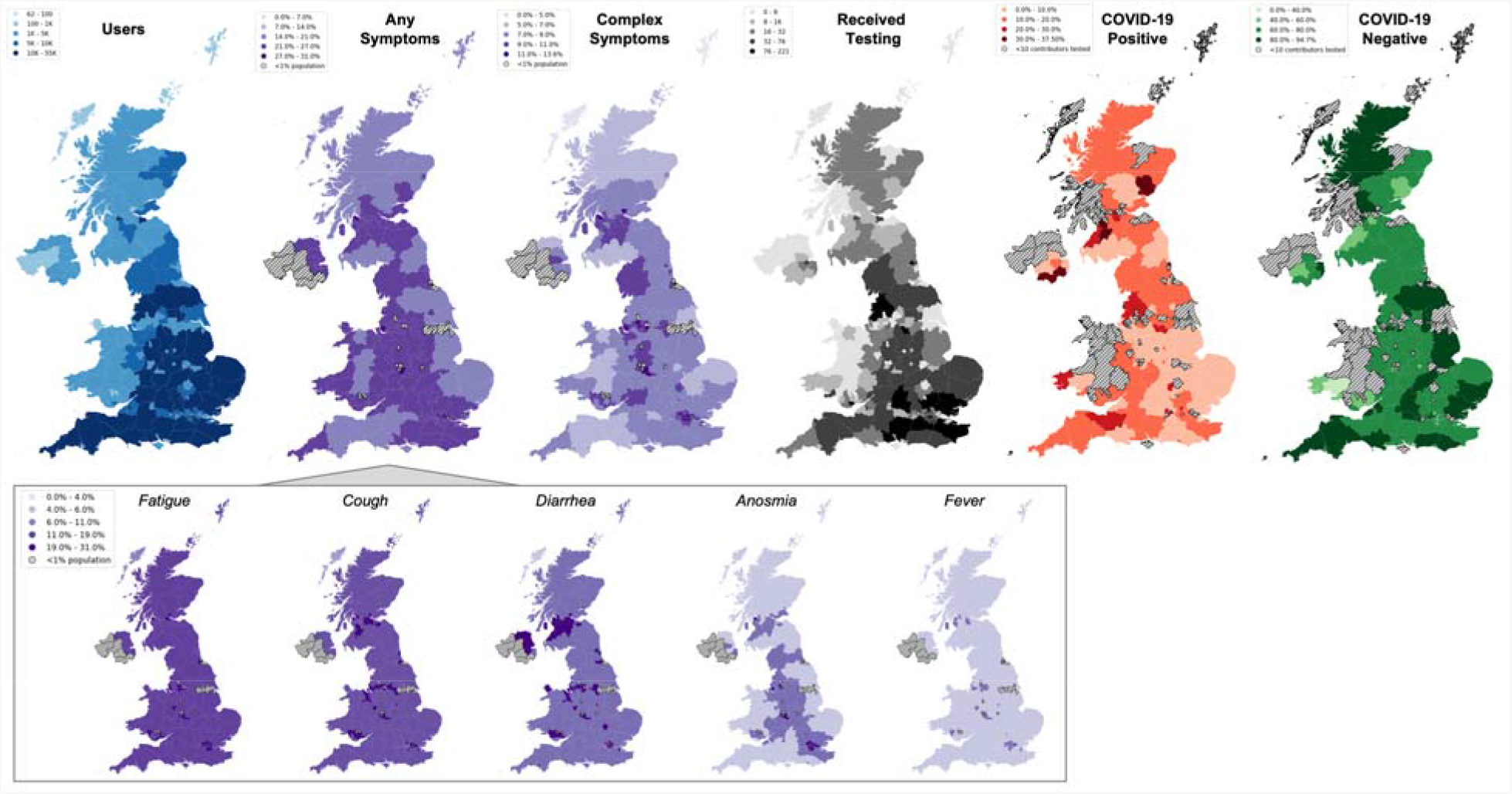
COVID Symptom Tracker use, reported symptoms, and testing results according to geographic location in the United Kingdom. Between March 24 and March 29, 2020, more than 1.6 million unique individuals downloaded the application and shared clinical and demographic information, as well daily symptoms and high-intensity occupational exposures across the United Kingdom (UK) (*blue map*). Population density of those presenting with any symptoms varied according to region with widespread reports of fatigue, cough, and diarrhea, followed by anosmia, and relatively, infrequently, fever (*inlay*). Examining those individuals who reported complex symptoms, defined as having cough or fever and at least one other of diarrhea, anosmia, and fever, reveals areas of the UK in potential need for more testing. Among the subset of the population that reported receiving a COVID-19 test (*black map*), areas with larger proportions of positive tests (*orange map*) appear to coincide with areas with high proportions of their population reporting complex symptoms, whereas some areas with low-complex symptom prevalence have received higher rates of testing and consequently more negative tests (*green map*). This example of real-time visualization of data captured by the COVID Symptoms Tracker may assist public health and government officials in reallocating resources, identifying areas with unmet testing needs, and detect emerging hot spots earlier.

**Fig 3.**
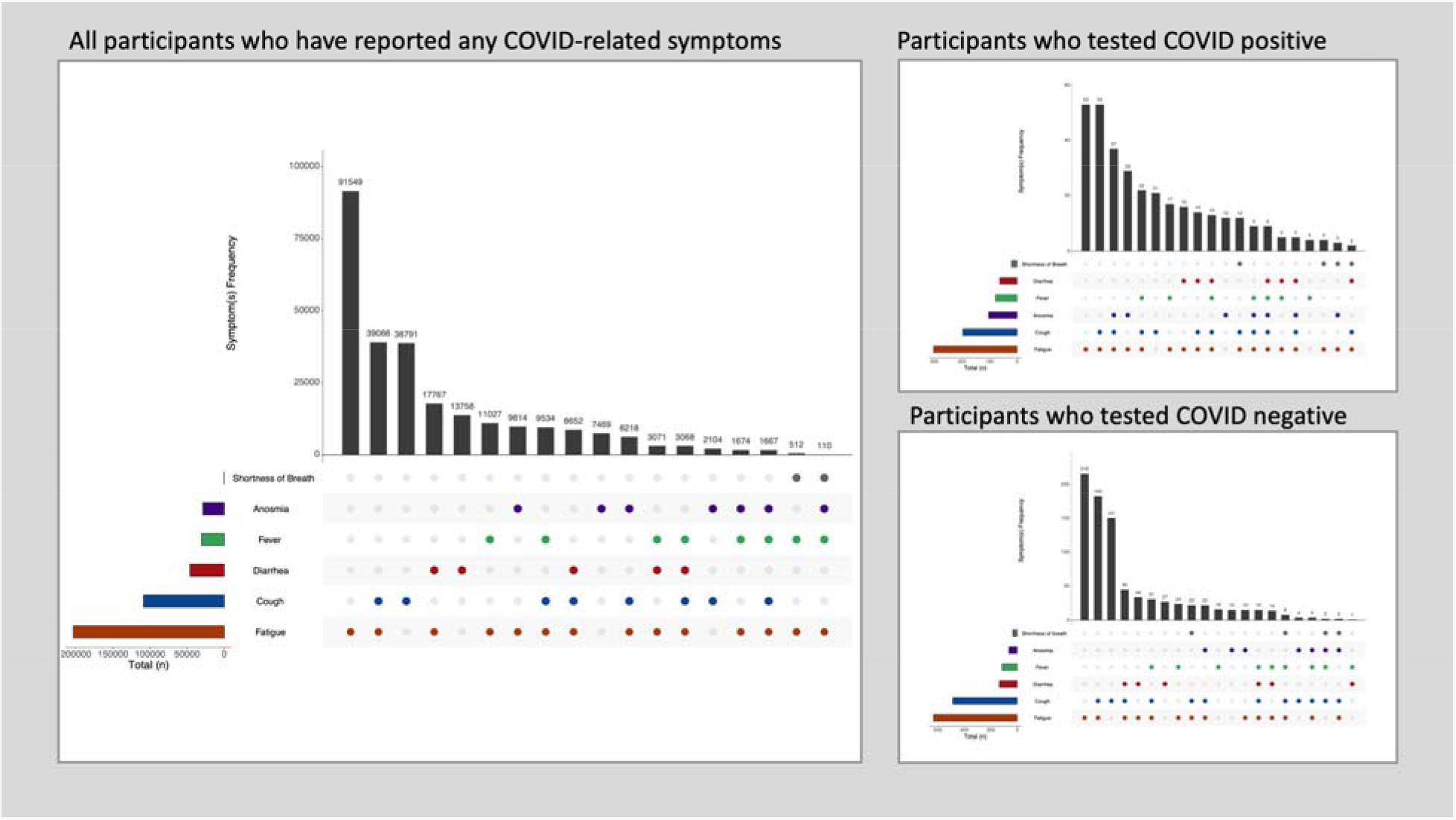
Symptoms reported through the COVID Symptom Tracker app. By March 27, 2020, 265,851 individuals reported any change in recent symptoms in the United Kingdom. (left). Participants provided data on whether they were tested for COVID-19 and the test result. 774 individuals reported having received a COVID-19 test (0.2% of those with symptoms). Symptom frequencies among those who tested COVID-19 positive (upper right) vs. negative (lower right)

With additional data collection, we will apply machine learning and other big-data approaches to identify novel patterns that emerge in dynamic settings of exposure, onset of symptoms, disease trajectory, and clinical outcomes. Our launch of the app within several large epidemiology cohorts that have previously gathered longitudinal data on lifestyle, diet and health factors and genetic information will allow investigation of a much broader range of putative risk factors to COVID-19 outcomes. With additional follow-up, we will also be uniquely positioned to investigate long-term outcomes of COVID-19, including mental health, disability, mortality, and financial outcomes. Mobile technology can also supplement recently launched clinical trials or biobanking protocols already embedded within clinical settings. For example, at the Massachusetts General Hospital (MGH) and Brigham and Women’s Hospital, we are deploying the tool within several ongoing clinical studies, centralized biobanking efforts, and healthcare worker surveillance programs. Healthcare workers are a particularly vulnerable population to COVID-19’s effects beyond infection, including work hazards from PPE shortages, emotional stress, and absenteeism. Real-time data generation focused within these populations will be critical to optimally allocate resources to protect our healthcare workforce and assess their efficacy.

In the near future, we hope to release our app as fair-use open source software to allow for its translation and development in other regions. We have also developed a practical toolkit for clinical researchers to facilitate local Institutional Review Board (IRB) and regulatory approval to facilitate use within research studies (www.monganinstitute.org/cope-consortium). This toolkit includes full detail of the questions asked within the app, consent documents, privacy policies, and terms of use for the mobile app. We believe our novel approach has demonstrated critical proof-of-concept for rapid repurposing of existing data collection approaches to implement scalable real-time collection of population-level data during a fast-moving global health crisis and National Emergency. We call upon our colleagues to work with us so that we may deploy all the tools at our disposal to address this unprecedented public health challenge.

## Data Availability

We welcome anyone who would like access to this growing dataset to join the COPE Consortium to gain access to the data following completion of a data use agreement. Details with how to become a member can be found at http://www.monganinstitute.org/cope-consortium.

## Notes

Use of the COVID-19 app in cohorts and informed consent as described was approved by the Partners Human Research Committee (Protocol #2020P000909) and is registered on ClinicalTrials.gov as NCT04331509.

## Acknowledgments

None.

## Funding

ATC is the Stuart and Suzanne Steele MGH Research Scholar. Development of the COVID Symptom Tracker mobile app was supported in kind by Zoe Global Limited and grants from the Wellcome Trust (212904/Z/18/Z).

## Author contributions

DAD and LHN drafted the manuscript. All authors contributed to the conceptualization, methodology, formal analysis, investigation, resources, data curation, and review and editing of the manuscript. ATC and TDS supervised the study and acquired funding;

## Competing interests

TDS is a consultant to Zoe Global Ltd. JW is an employee of Zoe Global Ltd. DAD and ATC previously served as investigators on a clinical trial of diet and lifestyle using a distinct mobile application that was supported by Zoe Global Ltd. Other authors have no conflict of interest to declare.

## Supplementary Materials

None.

## Notes

### Clinical Trial

NCT04331509

### Clinical Protocols

http://www.monganinstitute/cope-consortium

## References and Notes

1. M. Lipsitch, D. L. Swerdlow, L. Finelli, Defining the Epidemiology of Covid-19 — Studies Needed. New England Journal of Medicine 382, 1194–1196 (2020).

2. G. A. FitzGerald, Misguided drug advice for COVID-19. Science 367, 1434 (2020).

3. D. Wang et al., Clinical Characteristics of 138 Hospitalized Patients With 2019 Novel Coronavirus-Infected Pneumonia in Wuhan, China. JAMA, (2020).

4. C. Wu et al., Risk Factors Associated With Acute Respiratory Distress Syndrome and Death in Patients With Coronavirus Disease 2019 Pneumonia in Wuhan, China. JAMA Intern Med, (2020).

5. F. Zhou et al., Clinical course and risk factors for mortality of adult inpatients with COVID-19 in Wuhan, China: a retrospective cohort study. Lancet 395, 1054–1062 (2020).

6. W. J. Guan et al., Clinical Characteristics of Coronavirus Disease 2019 in China. N Engl J Med, (2020).

7. X. Yang et al., Clinical course and outcomes of critically ill patients with SARS-CoV-2 pneumonia in Wuhan, China: a single-centered, retrospective, observational study. Lancet Respir Med, (2020).

8. L. Fang, G. Karakiulakis, M. Roth, Are patients with hypertension and diabetes mellitus at increased risk for COVID-19 infection? Lancet Respir Med, (2020).

9. M. Day, Covid-19: ibuprofen should not be used for managing symptoms, say doctors and scientists. BMJ 368, m1086 (2020).

10. N. Chen et al., Epidemiological and clinical characteristics of 99 cases of 2019 novel coronavirus pneumonia in Wuhan, China: a descriptive study. Lancet 395, 507–513 (2020).

11. C. Huang et al., Clinical features of patients infected with 2019 novel coronavirus in Wuhan, China. Lancet 395, 497–506 (2020).

12. X. W. Xu et al., Clinical findings in a group of patients infected with the 2019 novel coronavirus (SARS-Cov-2) outside of Wuhan, China: retrospective case series. BMJ 368, m606 (2020).

13. P. J. Lyons, Coronavirus Briefing: What Happened Today in The New York Times. (New York, New York, 2020).

14. L. Pan et al., Clinical characteristics of COVID-19 patients with digestive symptoms in Hubei, China: a descriptive, cross-sectional, multicenter study. Am J Gastroenterol [Epub ahead of print] (2020).

15. J. S. Brownstein, C. C. Freifeld, L. C. Madoff, Digital disease detection--harnessing the Web for public health surveillance. N Engl J Med 360, 2153–2155, 2157 (2009).

16. B. Xu, M. U. G. Kraemer, C.-D. C. G. Open, Open access epidemiological data from the COVID-19 outbreak. Lancet Infect Dis, (2020).

